# Study on horizon-scanning with a focus on the development of AI-based medical products: citation network analysis

**DOI:** 10.1101/2021.02.25.21252496

**Authors:** Takuya Takata, Hajime Sasaki, Hiroko Yamano, Masashi Honma, Mayumi Shikano

## Abstract

**Objectives:** Horizon-scanning for innovative technologies that might be applied to medical products and require new assessment approaches/regulations will help to prepare regulators, allowing earlier access to the product for patients and an improved benefit/risk ratio. In this study, we focused on the field of AI-based medical image analysis as a retrospective example of medical devices, where many products have recently been developed and applied. We proposed and validated horizon-scanning using citation network analysis and text mining for bibliographic information analysis.

**Methods and analysis:** Research papers for citation network analysis which contain “convolutional*” OR “machine-learning” OR “deep-learning” were obtained from Science Citation Index Expanded (SCI-expanded) in the Web of Science (WoS). The citation network among those papers was converted into an unweighted network with papers as nodes and citation relationships as links. The network was then divided into clusters using the topological clustering method and the characteristics of each cluster were confirmed by extracting a summary of frequently cited academic papers, and the characteristic keywords, in the cluster.

**Results:** We classified 119,553 publications obtained from SCI and grouped them into 36 clusters. Hence, it was possible to understand the academic landscape of AI applications. The key articles on AI-based medical image analysis were included in one or two clusters, suggesting that clusters specific to the technology were appropriately formed. Based on the average publication year of the constituent papers of each cluster, we tracked recent research trends. It was also suggested that significant research progress would be detected as a quick increase in constituent papers and the number of citations of hub papers in the cluster.

**Conclusion:** We validated that citation network analysis applies to the horizon-scanning of innovative medical devices and demonstrated that AI-based electrocardiograms and electroencephalograms can lead to the development of innovative products.

**Article Summary:** *Strengths and limitations of this study:*

- Citation network analysis can provide an academic landscape in the investigated research field, based on the citation relationship of research papers and objective information, such as characteristic keywords and publication year.
- It might be possible to detect possible significant research progress and the emergence of new research areas through analysis every several months.
- It is important to confirm the opinions of experts in this area when evaluating the results of the analysis.
- Information on patents and clinical trials for this analysis is currently unavailable.

## INTRODUCTION

The application of innovative technologies to the development of medical products is expected to be a potential new treatment or diagnostic tool for diseases currently lacking these. Conversely, there may be cases where the application of conventional development and evaluation concepts and/or regulatory frameworks to innovative technologies is inappropriate. Therefore, the early identification of innovative technologies with a potential application to medical products through horizon-scanning would encourage regulatory authorities to establish new approaches to assess their quality, efficacy, and safety to advise developers and revise their regulations if necessary. This will contribute to timely patient access and improve the benefit/risk ratio of product ^1^.

The International Coalition of Medicines Regulatory Authorities (ICMRA), consisting of regulatory authorities from 30 countries and regions, has recognised the need to respond quickly to innovative technologies and shares the importance of ‘horizon-scanning’ to identify such technologies ^2^ among member authorities. The ICMRA Innovation concept note ^3^ describes horizon-scanning as a broad-reaching information-gathering and monitoring activity to anticipate emerging products and technologies and potentially disruptive research avenues. Traditionally, horizon-scanning has been predominantly conducted in Europe for policy-making, scientific research funding, and health care budgeting purposes, by surveying a variety of sources — such as the Internet, government, international organisations and companies, databases, and journals — using the Delphi method, for example ^4 5^. Recently, the European Commission(EC) has published some reports including “Weak signals in Science and Technologies 2019 Report based on Tools for Innovation Monitoring (TIM) ^6^, which uses text mining and keywords in the scientific literature. The Japanese National Institute of Science and Technology Policy (NISTEP) also uses the Delphi method and a digital tool to analyse academic papers with the top 1 % of citations to contribute to science and innovation policy planning.

Hines et al. reported that, in the medical and health care field, the majority of horizon-scanning methods used were manual or semiautomated, with relatively few automated aspects; this may be resolved in the not-too-distant future via the rapidly evolving fields of machine learning and artificial intelligence ^5^.

It is nearly impossible to understand the whole picture of the extremely large and fragmented results of research and technological development and the limitations of existing methods are now being pointed out in many fields.

To solve this challenge, a computer-based approach can be used to complement the expert-based approach as it fits the scale of the information (Börner et al. 2003; Boyack et al. 2005). In particular, the citation-based approach assumes that the cited papers and research topics of the cited papers are similar. Analysing this citation network allows us to understand the structure of the research areas constituting the large volume of papers we are able to read. These methods have been widely used as powerful tools to visualise and understand the structure of a research field and to identify new trends and research directions; they have been proven effective through various studies (Chen 1999 ^7^; Chen, Cribbin & al., 2003 ^8^; Small, 1999 ^9^).

For example, Kajikawa et al. (2007) ^10^ used citation network analysis to effectively and efficiently track emerging research areas in the field of sustainable science. Similar approaches have been applied to a wide range of fields, including energy research (Kajikawa et al., 2008 ^11^), regenerative medicine (Shibata et al., 2011 ^12^), robotics, and gerontology (Ittipanuvat et al., 2014 ^13^). Sakata et al. (2012) ^14^ proposed a meta-structure of academic knowledge on patent and innovation research to effective assist policy discussions for intellectual property system reform. Using a citation-based approach, this study analyses the academic landscape of patent and innovation research to understand the current structure and trends of research and to detect major sub-research fields and core papers within it. They have shown that network analysis and machine learning methods are useful for understanding and predicting the development of technologies such as solar cells ^15^ and nanocarbon ^16^. Citation network analysis and text mining are useful tools for R&D strategists and policymakers in many fields to understand the broad scope of scientific and technological research and make decisions for worthwhile investments in promising technologies.

This paper proposes and discusses a methodology for horizon-scanning to identify innovative technologies that may be applied to medical products by utilising citation network analysis methods and text mining. In this paper, we focus on AI-based medical image analysis as a retrospective example of AI-based medical devices that have been developed in recent years, applied in many fields, and selected for consideration in ICMRA1. By analysing research papers on the development of AI-based medical technologies, we explored the network-like characteristics of this field and proposed a prediction procedure for innovative technologies related to the medical field.

## METHODS

### Extraction of paper data for analysis

To track the development history of AI-based medical image analysis and to select keywords for the extraction of the papers for citation network analysis, we selected 13 key articles ^17–27^ (presented in Table 1), including several papers cited in the review article ^28^ on the application of deep learning in medical image analysis and a study ^29^ that lead to the clinical development of IDx-DR, a retinal imaging software approved as a medical device by the US FDA in 2018.

**Table 1.**
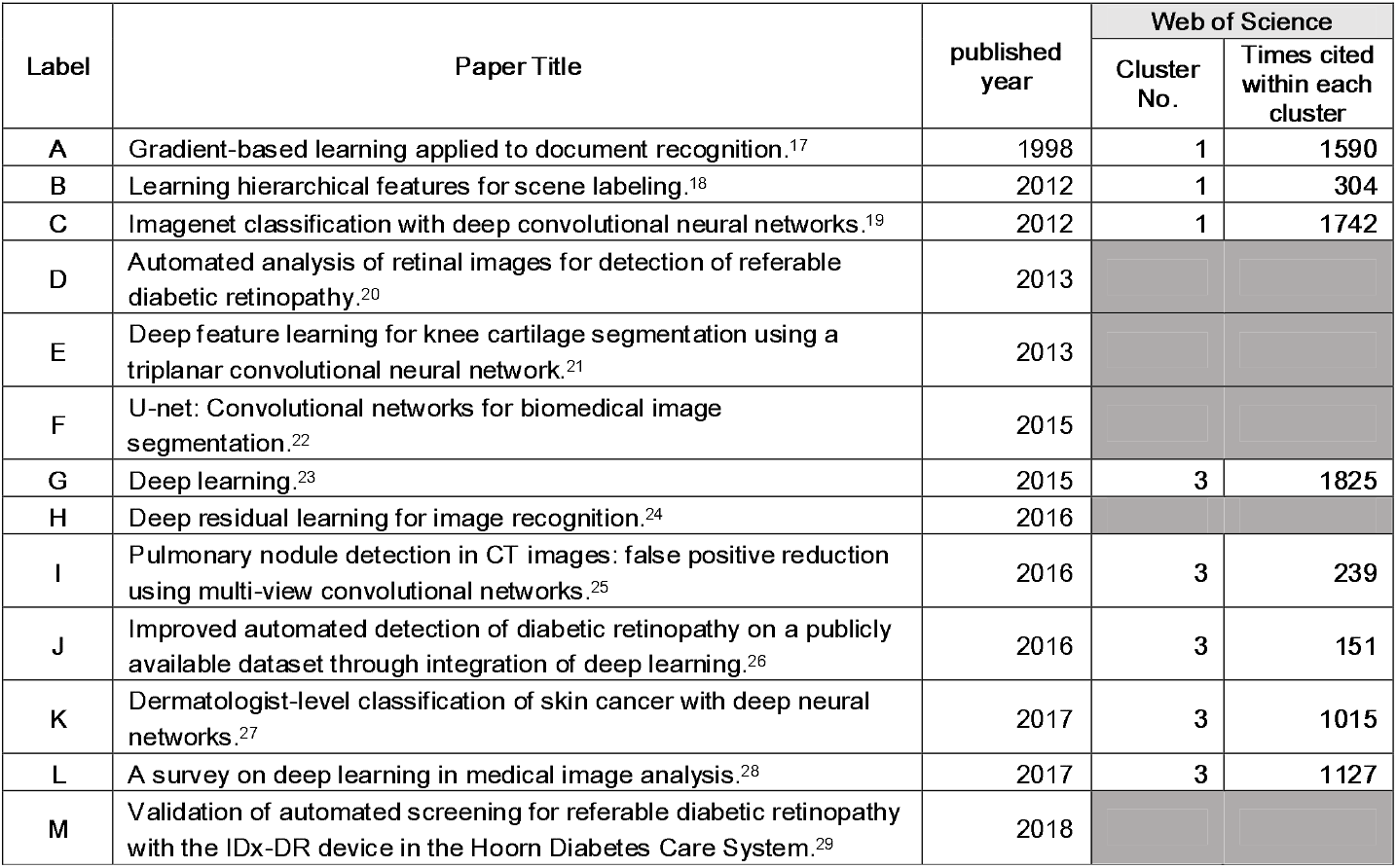
Key articles and the clusters in which they are contained. The key articles that have contributed to the development of AI-based medical image analysis were selected based on a review article on AI-based medical image analysis ^28^. The clusters obtained from the citation network analysis of these articles are indicated. The clusters are numbered in descending order of the number of constituent papers included. The cells for papers not included in the analysis were shadowed.

In addition to the query setting “convolutional” OR “deep learning” in the review article of medical image analysis (G Litjens et al, 2017) ^12^, we used “machine-learning” to include a wide range of conventional studies. As a result, we obtained 140,794 papers that contain “convolutional*” OR “machine-learning” OR “deep-learning” from the Web of Science Core Collection (WoS, Thomson Reuters), between 1 January 1900 and 31 December 2020.

For analysis, data sets were created between 1 January 1900 and 31 December 2012 to 2019 and the cluster that contains key articles for each year was identified.

### Citation network analysis

In this study, the citation network was converted into an unweighted network with papers as nodes and citation relationships as links. Papers with no citations as the largest component were considered digressional and were ignored in this study (Step 2 in Fig 1). The core paper with the highest number of citations is located at the centre of the citation relations. Papers with no citation relationship with other papers were considered deviant and ignored in this study. The network is then divided into several clusters using the topological clustering method. Topological clustering is a clustering method based on the graph structure of a network; here, we use modularity maximisation. A cluster is a module in a citation network and is a group of papers in which the citation relations are divided using a modularity (Q value) maximisation method and are densely aggregated (Louvain method) ^16 30^. The modularity maximisation method appreciates network partitioning such that the intracluster is dense and the intercluster is sparse. The modularity maximisation method determines an optimal partitioning pattern by extracting the partitioning pattern that maximises the modularity using a greedy algorithm. Q is an evaluation function of the degree of coupling within a cluster and between clusters, as follows:

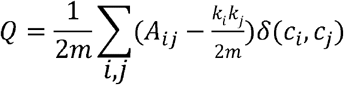

where *A*_*ij*_ represents the weight of the edge between *i* and *j, k*_*i*_ = ∑_*j*_ *A*_*ij*_ is the sum of the weights of the edges attached to vertex i, c_i_ is the community to which vertex i is assigned, δ-function δ (u, v) is 1 if *u*= *v* and 0 otherwise, and 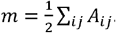

**Fig 1.**
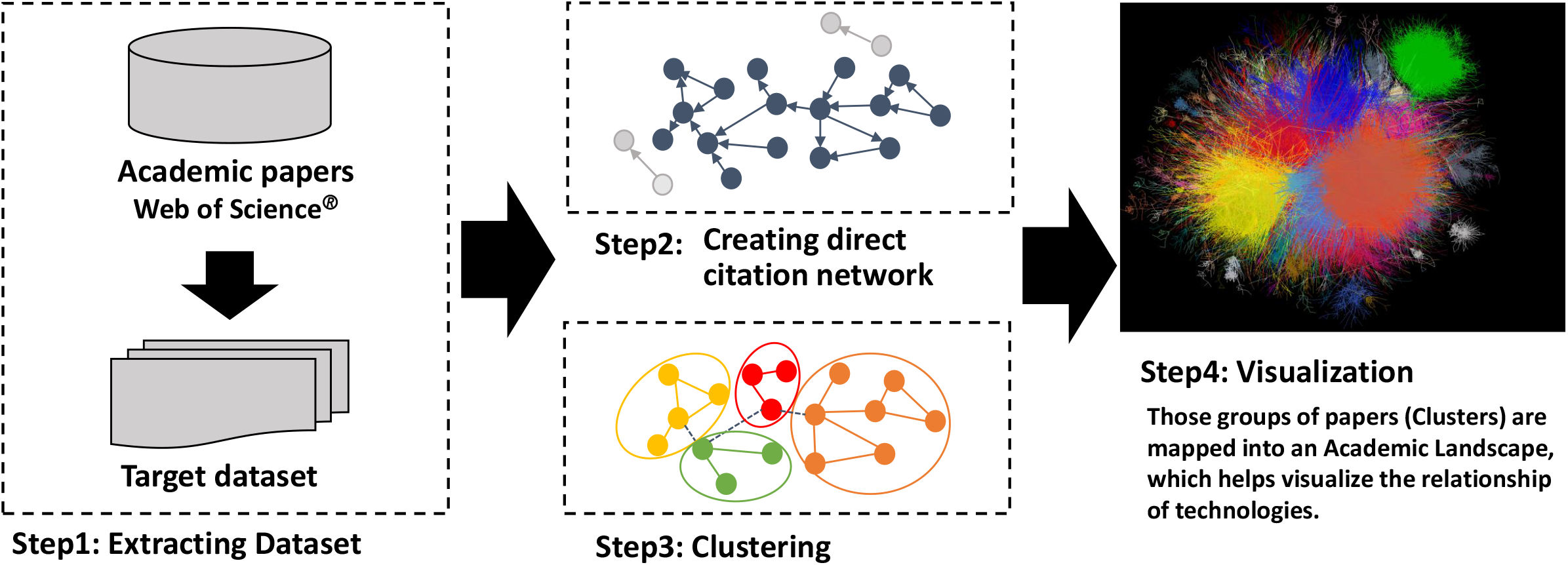
Steps of clustering and making Academic Landscape based on citation network ^31 32^. This Figure has been published in reference 11. The procedure of the citation network is as follows: extraction dataset of academic papers for analysis (Step 1).

The clusters are assigned labels corresponding to the size of the number of papers included. The characteristics of each cluster were confirmed by extracting a summary of frequently cited academic papers in the cluster and the characteristic keywords in the cluster.

In addition, we computed the term frequency-inverse cluster frequency (TF-ICF) to extract the characteristic keywords of each cluster. The term frequency TF gives a measure of the importance of a term in a particular sentence. The inverse cluster frequency ICF provides a measure of the general importance of a term. The TFICF of a given term i in a given cluster j is given by:

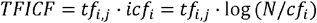

where N is the total number of sentences. Each cluster was labelled based on the resulting keywords and sentences.

To confirm the trends in the research field, we extracted the mean or median year of publication of papers in each cluster, as well as information on journals, authors, and affiliated institutions.

After clustering the network, visualisation is converted to intuitively infer a relation among these clusters. We use a large graph layout (LGL) based on a force-direct layout algorithm (Adai et al., 2004) ^31 32^. This layout can display the largest connected component of the network to generate coordinates for nodes in two dimensions. We visualise the citation network by expressing inter-cluster links with the same colour (Step 4 in Fig 1). However, the position of the clusters and the distance between clusters do not indicate an approximation of the content. An overview of this is shown in Fig 1.

For the extracted dataset, the citation network was converted into an unweighted network with papers as nodes and citation relationships as links (Step 2). The network was then divided into several clusters using the topological clustering method (Step 3). In addition, a large graph layout (LGL) – based on a force-direct layout algorithm – displayed the largest connected component of the network to generate coordinates for the nodes in two dimensions, visualising the citation network by expressing inter-cluster links with the same colour (Step 4).

## RESULTS

### Results of citation network analysis

We analysed the citation network of 140,794 papers and found that 119,553 (85 %) formed a citation network; this was divided into 36 clusters by extracting the largest linkage component from all linkage components via direct citation of papers (excluding the grey linkage not involved in cluster formation shown in Fig 1 and 2). The contents of the top 10 clusters, which contain approximately 75 % of the papers in a citation network, were estimated from the characteristic keywords appearing in each cluster and the titles and abstracts of the papers with the highest number of citations. The cluster numbers (number of papers) and their contents are listed below.

**Fig 2.**
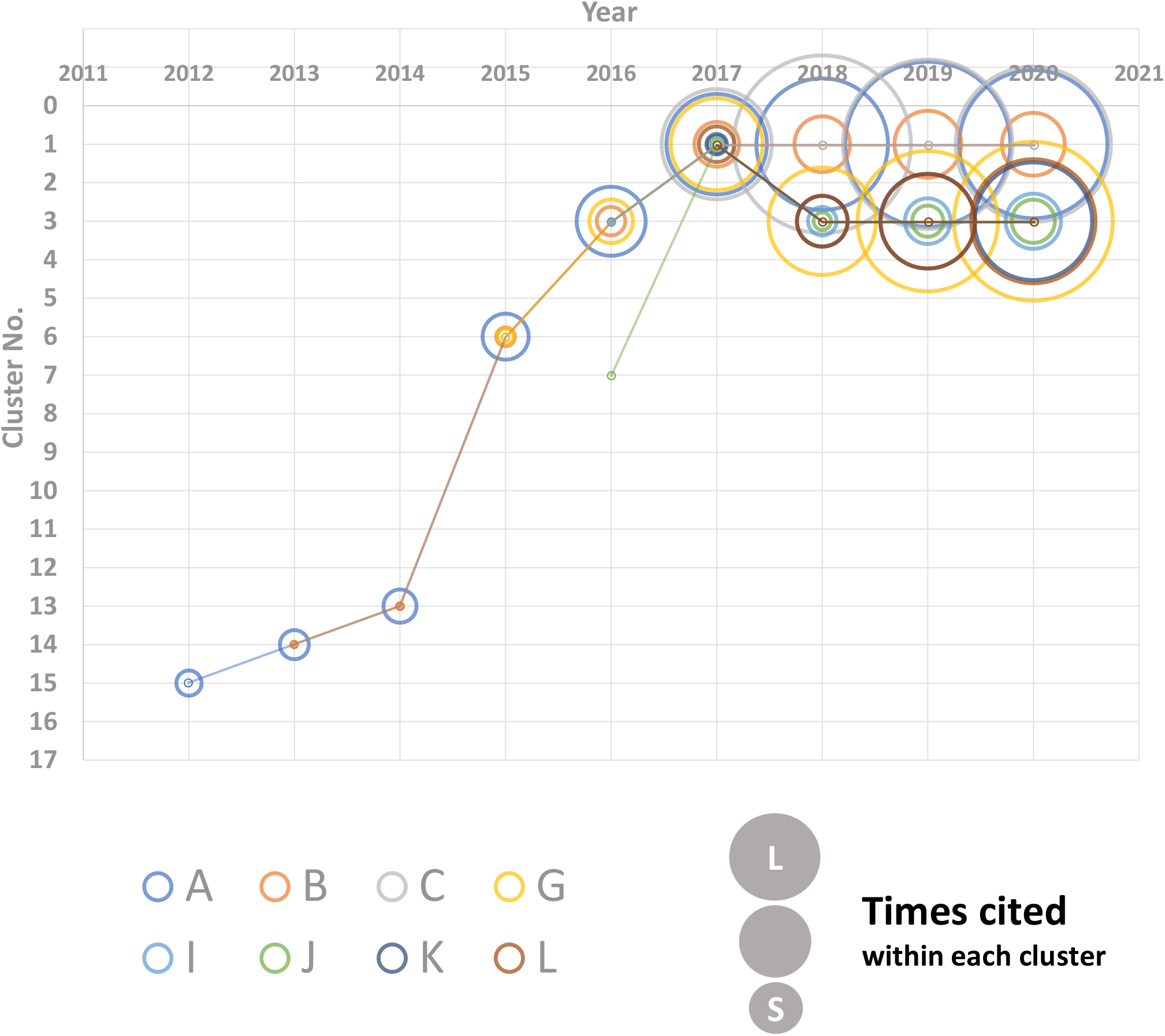
Tracking clusters containing key articles. Papers obtained from WoS published up to the indicated year were analysed. The cluster numbers that contained the eight key articles shown in Table1 were plotted and the size of the circles represents the approximate number of citations in the cluster for each paper.

Cluster 1 (11,711): basic studies on deep learning and convolutional neural networks (CNNs), including geographic information system (GIS) image analysis using remote sensing.

Cluster 2 (7,597): drug discovery technologies related to proteins, peptides, *etc*., using machine learning.

Cluster 3 (5,323): applied research in medical image analysis.

Cluster 4 (4,340): feature classification using ensemble methods to increase accuracy by combination.

Cluster 5 (3,665): natural language processing of clinical records.

Cluster 6 (2,691): application of deep learning to fault diagnosis, for example, motor condition monitoring for machines running on electric motors.

Cluster 7 (2,497): machine learning (ML) and data mining (DM) methods for cyber analysis.

Cluster 8 (2,311): application to traffic flow information analysis for the implementation of intelligent transport systems.

Cluster 9 (2,281): single-image super-resolution (SR) to reconstruct high-quality data.

Cluster 10 (2,232): classification of individuals based on the analysis of text information from social media, such as emotions and behaviour.

Table 1 presents the clusters in which the key articles were included. Three papers (A, B, C) based on image recognition are found in cluster 1 and five on image diagnosis in cluster 3, including the review article “Deep Learning” ^23^, which is often cited in medical field papers. This indicates that clusters related to medical imaging were appropriately formed in cluster 3.

### Tracking the time series of key articles

We analysed papers published each year and identified the cluster containing the key papers in Table 1 and the number of citations within the cluster to assess the position of the research on medical imaging in the past. As shown in Fig. 2, all the papers were included in the same cluster until 2015 and the rank of cluster number increased by one until 2014. In 2015, the number of papers in this field increased rapidly and the rank of cluster number rose from 13th in 2014 to 6th, suggesting that great scientific attention has increased. In 2016, a key paper on the imaging diagnosis of diabetic retinopathy (J in Table 1) was in cluster 7, which comprised papers on medical image analysis, and the other seven key articles were in cluster 3. Subsequently, in 2017, cluster 1 contained all of the key articles but, from 2018 onwards, a new separate cluster containing papers on image analysis using deep learning was formed; it can be seen that the number of citations of the key articles also increased.

Thus, most of the key articles were in one or two clusters, suggesting that the clusters related to the targeted AI-based medical image analysis were properly formed. The research status of the clusters can also be confirmed by the cluster numbers, which reflect the number of papers comprising the cluster and the number of citations of the key articles.

### Recent research trends in AI-based medical products

To detect the latest research trends in AI-based medical products, we focused on ‘younger’ clusters with an average publication year later than 2017 as research progress could be observed over three years for AI-based medical image analysis (Fig 3). We re-analysed clusters 3, 15, 12, 5, 13, and 2, which were considered to be closely related to AI-based medical technologies. Clusters 3, 15, 12, 5, 13, and 2 were listed in order of average publication year. Table 2 lists the sub-clusters formed by re-analysis of the most cited article ^23 33–64^ (hub-paper) in each subcluster, suggesting recent research trends in this field as follows. Cluster 3: applied research in medical image analysis. Cluster 15: electrocardiogram, electroencephalogram, and other electrical biosignals of human activity. Cluster 12: human activity recognition. Cluster 5: natural language processing of clinical records. Cluster 13: neuroimaging analysis. Cluster 2: drug discovery with machine learning related to proteins, peptides, *etc.*

**Table 2.**
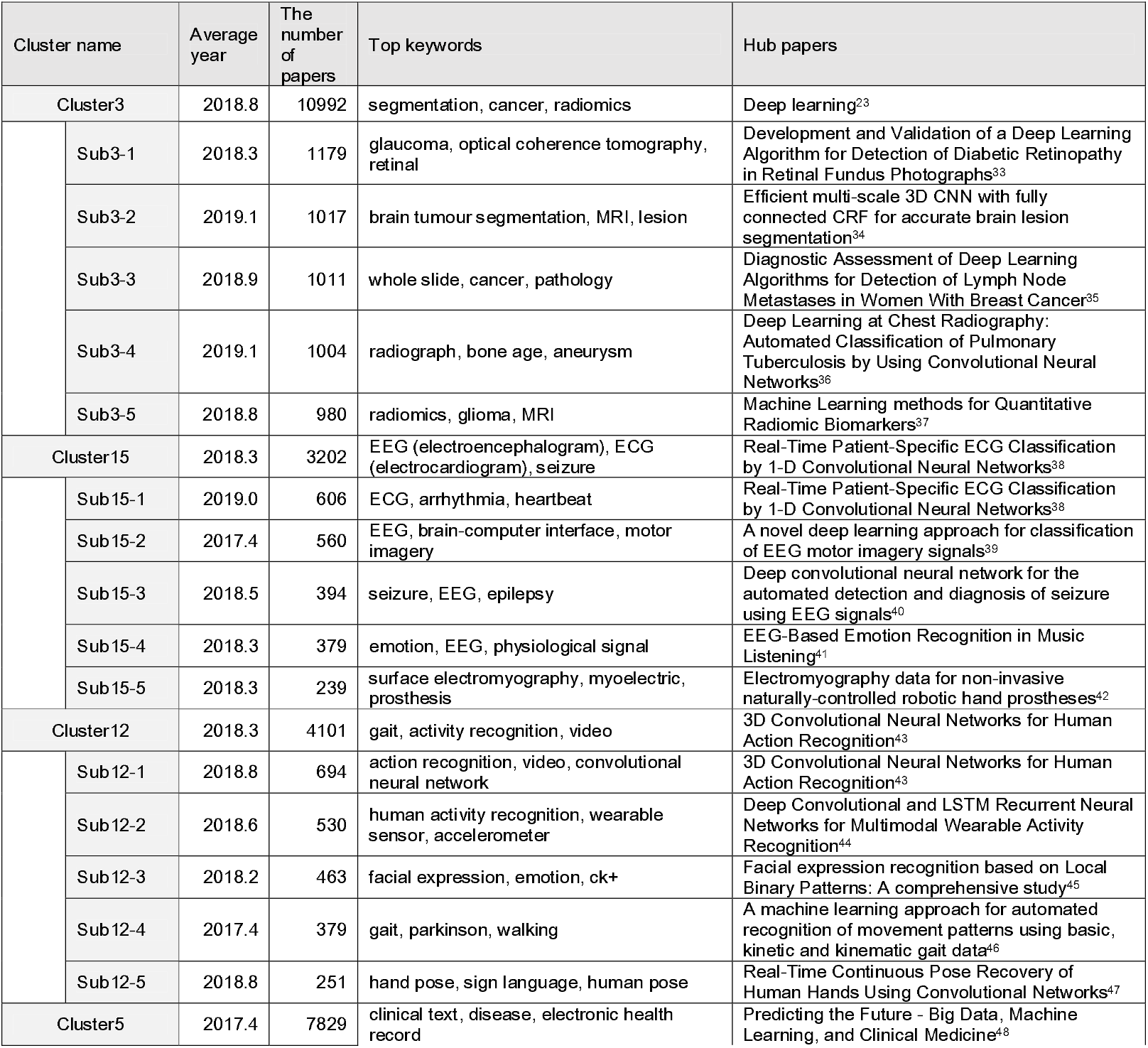
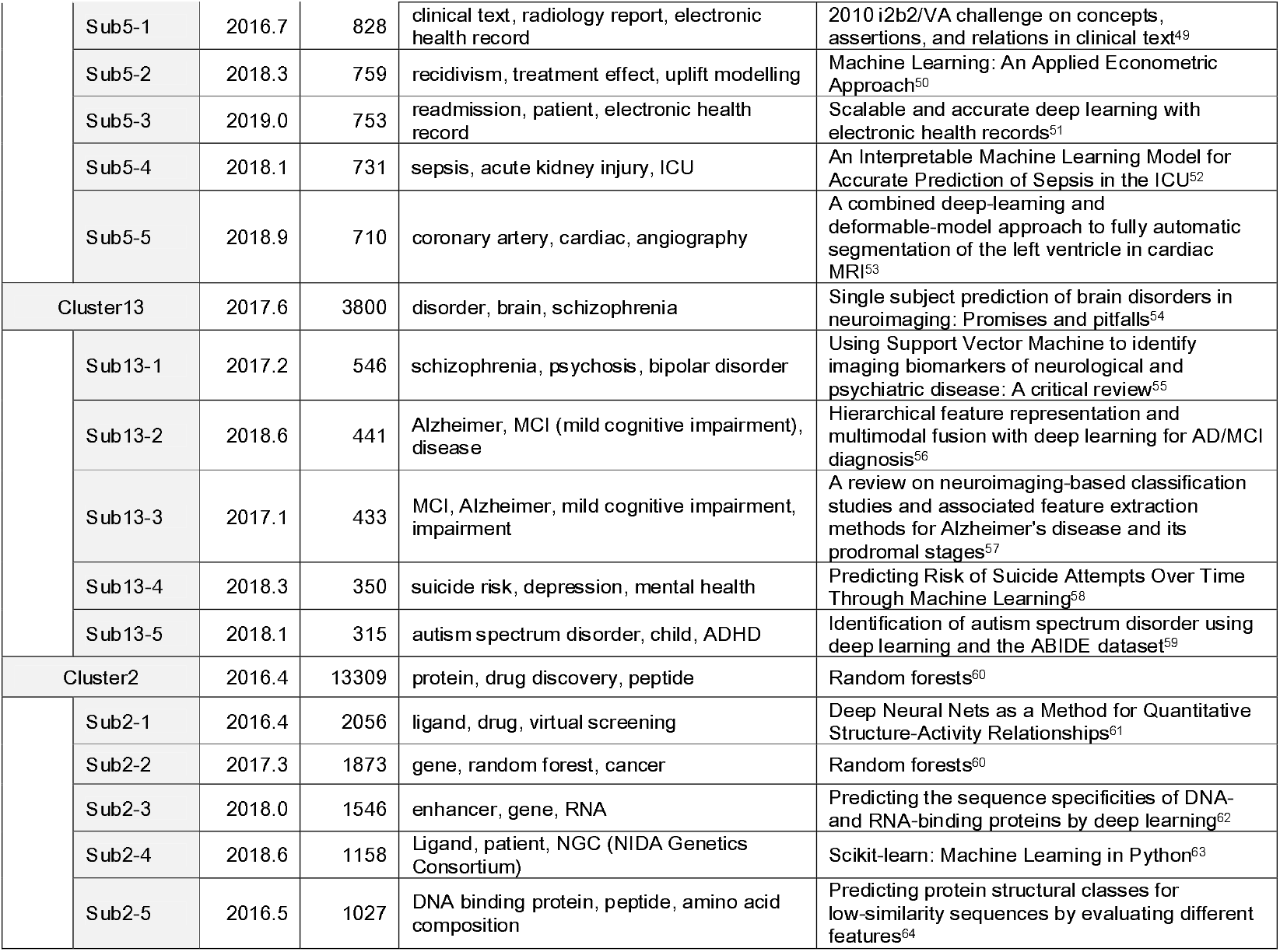
Sub-clustering results for clusters of AI-based medical technologies. The clusters of AI-medical technologies were re-analysed and the characteristics of the top five sub-clusters, that is, the number and average of publications of constituent papers, specific keywords, and the title of hub-paper are shown.

**Fig 3.**
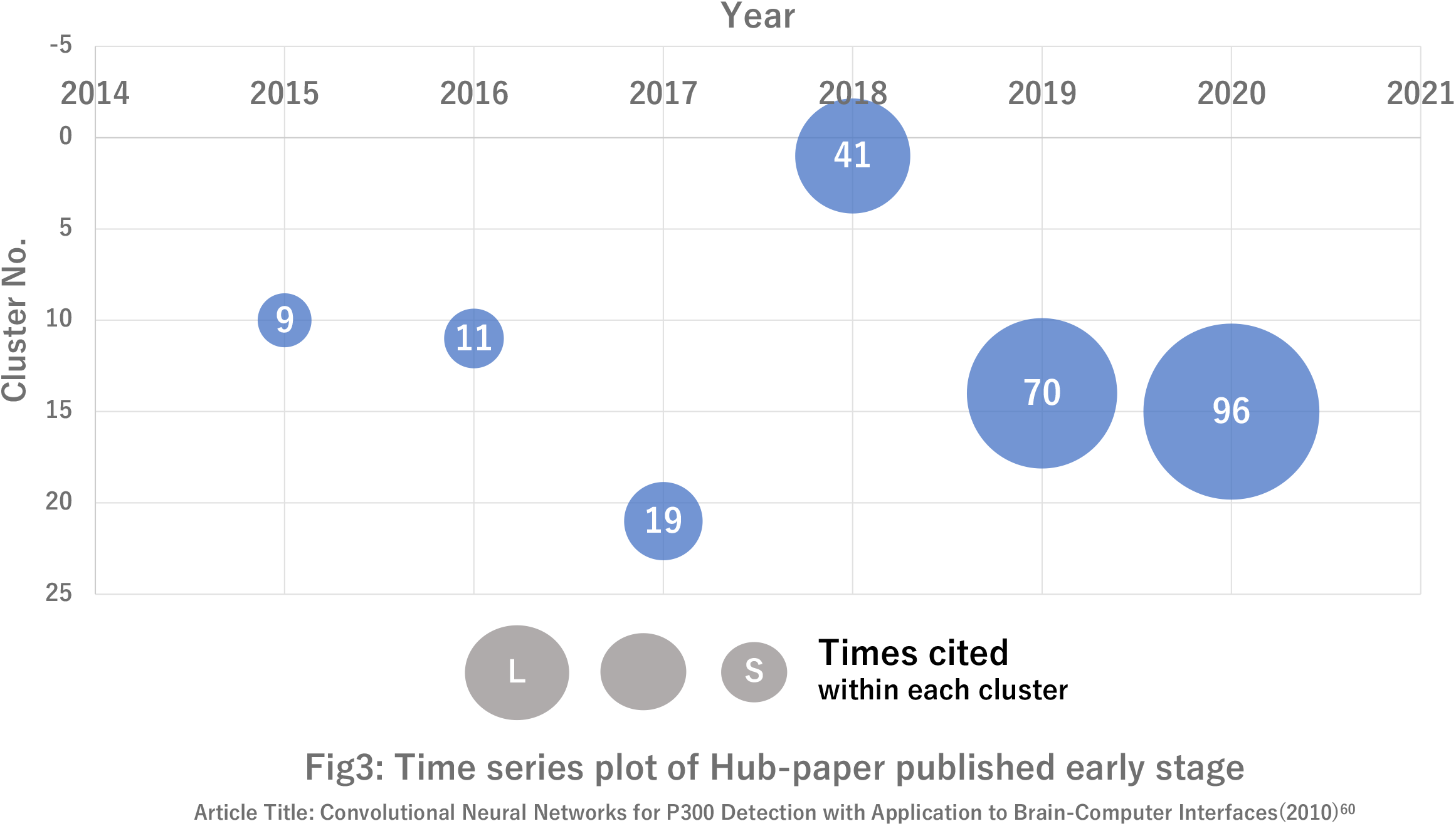
Tracking clusters related to ECG and EEG. Papers obtained from WoS published up to the indicated year were analysed. Clusters on ECG and EEG were first detected in 2015. The cluster on the ECG and EEG is indicated as a cluster number. The size of the circles indicates the approximate citation frequency of key article ^60^ and the number in each circle represents the number of citations in the cluster.

Among these AI-based medical technologies, EEG analysis was identified for applications in epileptic seizure prediction, emotional analysis, and brain-computer interfaces, for which the FDA issued draft guidance on non-clinical and clinical trials in 2019.

Electrocardiograms (ECGs) and electroencephalograms (EEGs) in cluster 15 are most likely to be applied to new medical devices; therefore, we tried to follow the cluster containing a key paper on the application of deep learning to EEG analysis by Cecotti & Graser(2010) ^65^, which was one of the triggers for the development of this field. During 2015 – 2016, the article was included in the same cluster as other neuroimaging techniques, such as MRI (MEG, fNIRS, etc.). In 2017, the key article was found in a separate cluster numbered 20 from other neuroimaging techniques, suggesting a new cluster specific to the application of deep learning to EEG was formed. Then, in 2018, the article was included in cluster 1 of the applications of deep learning in various fields but was included in a specific cluster re-formed, numbered 14 and 15 in 2019 and 2020, respectively, and the number of citations of the article increased. This suggests that research in this field has developed rapidly since 2017.

## DISCUSSION

The development of medical products based on innovative technologies may sometimes not be amenable to current development and evaluation approaches or regulatory frameworks. Horizon-scanning for such technologies will contribute to earlier access to the product for patients and a better benefit/risk ratio for the product by encouraging regulatory authorities to develop new guidance/regulations. Experts who have a deep understanding of innovative technologies would be able to predict the development of medical products based on the technology but experts on all evolving innovative technologies may not be available to regulatory authorities. However, it might sometimes be inappropriate to narrow the scope of consideration based solely on expert’s opinions, as information from experts is subjective and the outcome depends upon the choice of the expert ^66^. Therefore, it is more efficient and appropriate to use a method based on objective information as a primary screening tool for horizon-scanning to identify candidate technologies that may require new guidance or regulation. In this study, we have shown that citation network analysis and text mining are suitable for this purpose. We used these methods to classify a large number of papers in the field by research topic and identified the topics of the clusters based on the characteristic keywords of the clusters and the titles of the most cited papers. We also objectively evaluated the attention and novelty of the topic based on the number of papers and the median year of publication.

In this study, we examined the possibility of using this analysis method for horizon-scanning targeting AI-based medical image analysis as a typical example that requires new regulatory frameworks and evaluation approaches ^67^. IDx-DR, an image analysis software for the automatic diagnosis of diabetic retinopathy, received US FDA certification in 2018. The AI characteristics are self-learning, the algorithm for learning data during the development of a medical product is in a black box, and performance changes as the product continues learning during clinical use. This has become an interesting dilemma for regulators ^67^.

We assessed the feasibility of using citation network analysis and text mining to identify trend history in AI-based medical image analysis research and development, which is as follows: Research on convolutional neural networks (CNNs), the currently leading technology in deep learning arose in the 1970s, renewed interest in neural networks was Werbos’s multi-layer networks (1975) ^68^ and LeNet (1998) ^17^ – a CNN-based handwritten number recognition system – was developed and succeeded by a CNN called AlexNet (2012) ^19^, which was a key trigger for renewed interest in neural networks.

Later, the U-net ^22^ architecture was proposed, which consists of an upsampling section that uses “up” convolution to increase the image size. Furthermore, the combination of CNNs and recurrent neural networks (RNNs), represented by long short-term memory (LSTM), has been applied to analysis involving time-series data ^28 44^. We evaluated 13 key articles, including these milestones in the development of AI-based medical image analysis, to determine how key articles could be captured by citation network analysis and found that eight were identified in one or two clusters (Table 1), with a concentration of the characteristic keywords of the clusters, and the titles and abstracts of the articles with the highest number of citations confirmed that the clusters were related to AI-based medical image analysis and that it was possible to identify actual research trends. In addition, we analysed the papers reported each year and found that the number of constituent papers of the cluster containing the key articles increased dramatically after 2014, with the rank rising from 13th to 6th, suggesting that the technology related to diagnostic imaging has progressed dramatically. This might have led to a major clinical trial of IDx-DR in 2017. Since then, there has been a further increase in research activity in this field, as can be seen from the rank of cluster number and number of citations in the key articles. Five of the 13 selected articles were not included in the analysis: three papers were not included in the Web of Science Core Collection and the other two (MD Abràmoff et al. (2013) ^20^, AA van der Heijden et al. (2018) ^29^) on clinical evaluation were not found with the set query; this is because there was no mention of the underlying technology in the abstract or title, and the methods were mainly described as product names or computer detection in both papers.

Next, we explored recent trends in the development of new medical products using AI by re-analysing ‘young’ clusters with a late average of the publication year of constituent papers to identify more specific topics by sub-clustering (Table 2). We focused on EEG and ECG, which have the potential to lead to the development of new medical devices, and followed the cluster containing the key article on this topic. As shown in Fig 3, the increase in constituent papers and the number of citations of the key article suggested that this topic has made significant progress during 2017 – 2018, which might be related to the US FDA issuing draft guidance on brain-computer interfaces in 2019 ^69^.

This study also showed that analysis every several months might allow us to identify the candidate topics to further investigate through the rapid rise of the rank of cluster number, i.e., a sharp increase in constituent papers (2014 – 2015 in Fig. 2 and 2017 – 1018 in Fig 3), or the emergence of a new cluster spun out of the original one (2017 – 2018 in Fig. 2 and 2016 – 2017 in Fig 3), which may be a signal of significant research progress.

However, this analysis method has the following limitations: papers in major journals are included in WoS relatively quickly after publication but there might be a delay of approximately six months for almost all journals and some research areas may not be reflected in WoS quickly enough, which may delay the identification of research trends. Although data are not shown in this paper, we also analysed the papers obtained from PubMed; however, approximately 30 % of the papers formed a citation network and only 5 of the 13 key articles were included. One of the possible reasons for not being able to extract appropriate research papers from PubMed was that many of the papers did not use terminology related to AI-based technologies. This suggests that the choice of the literature database according to the target technology is also critical. Furthermore, research results in the field of machine learning, which covers basic technologies in the field of AI, are sometimes published in venues such as arXiv.com, where researchers can directly exchange papers with each other via the Internet; therefore, the latest results cannot be covered by databases of academic papers, such as WoS or PubMed.

Another possible bias, as mentioned by Takano et al. (2017) ^70^, is that researchers mainly check and cite papers written in their native language or journals they contribute to, or that they tend to search and cite papers using the same terminology and not others — even when the technological meaning is the same.

It is important to hear the opinions of experts in the field regarding the candidate topics to be investigated, which will result in overcoming the limitations described above.

It is expected that this citation network analysis will be established as a primary horizon-scanning method by continuing the study in other fields and organising the analysis conditions and points to be noted according to the characteristics of the target technology.

## Data Availability

The data that support the findings of this study are available from the corresponding author, Mayumi Shikano, upon reasonable request.

## Author Contributions

MS obtained developed the research design, interpreted the results. TT investigated literatures, analyzed the data and interpreted the results. HS and HY designed the methodology, software and interpreted the results. MH designed the data editing. TT and MS drafted manuscript. All authors have read and approved the final manuscript.

## Acknowledgment

We would like to thank Dr. Hidefumi Kobatake for his advice on the history of AI technology development. We also thank Dr. Rika Wakao, Dr. Masafumi Shimokawa, and Ms. Ai Fukaya for their help.

## References

1. ICMRA. Innovation Strategic Priority Project Report. 2019 [Available from: http://www.icmra.info/drupal/sites/default/files/2019-04/Innovation%20Strategic%20Priority%20Final%20Report.pdf2020.

2. ICMRA. Innovation | International Coalition of Medicines Regulatory Authorities (ICMRA). [Available from: http://www.icmra.info/drupal/en/strategicinitiatives/innovation accessed January 12 2021.

3. ICMRA. ICMRA Strategic Priority on Innovation Concept Notes 2017 [Available from: http://www.icmra.info/drupal/sites/default/files/2017-12/ICMRA%20Innovation%20Concept%20Note_0.pdf.

4. OECD. Overview of Methodologies [Available from: https://www.oecd.org/site/schoolingfortomorrowknowledgebase/futuresthinking/overviewofmethodologies.htm accessed January 6 2021.

5. Hines P, Yu LH, Guy RH, et al. Scanning the horizon: a systematic literature review of methodologies. Bmj Open 2019;9(5) doi: 10.1136/bmjopen-2018-026764

6. EU Science Hub - European Commission. ESH-E. Tools for Innovation Monitoring. 2017 [Available from: https://ec.europa.eu/jrc/en/scientific-tool/tools-innovation-monitoring.

7. Chen C. Visualising semantic spaces and author co-citation networks in digital libraries. Information processing & management 1999;35(3):401–20.

8. Chen C, Cribbin T, Macredie R, et al. Visualizing and tracking the growth of competing paradigms: Two case studies. Journal of the American Society for information Science and Technology 2002;53(8):678–89.

9. Small H. Visualizing science by citation mapping. Journal of the American society for Information Science 1999;50(9):799–813.

10. Kajikawa Y, Ohno J, Takeda Y, et al. Creating an academic landscape of sustainability science: an analysis of the citation network. Sustainability Science 2007;2(2):221–31.

11. Kajikawa Y, Yoshikawa J, Takeda Y, et al. Tracking emerging technologies in energy research: Toward a roadmap for sustainable energy. Technological forecasting and social change 2008;75(6):771–82.

12. Detecting emerging research fronts in regenerative medicine by citation network analysis of scientific publications. PICMET’09-2009 Portland International Conference on Management of Engineering & Technology; 2009. IEEE.

13. Ittipanuvat V, Fujita K, Sakata I, et al. Finding linkage between technology and social issue: A Literature Based Discovery approach. Journal of Engineering and Technology Management 2014;32:160–84.

14. Sakata I, Sasaki H, Kajikawa Y. Identifying knowledge structure of patent and innovation research. Journal of Intellectual Property Association of Japan 2012;8(2):56–67.

15. Sasaki H, Hara T, Sakata I. Identifying Emerging Research Related to Solar Cells Field using a Machine Learning Approach. Journal of Sustainable Development of Energy Water and Environment Systems-Jsdewes 2016;4(4):418–29. doi: 10.13044/j.sdewes.2016.04.0032

16. Sasaki H, Fugetsu B, Sakata I. Emerging Scientific Field Detection Using Citation Networks and Topic Models—A Case Study of the Nanocarbon Field. Applied System Innovation 2020;3(3):40.

17. Lecun Y, Bottou L, Bengio Y, et al. Gradient-based learning applied to document recognition. Proceedings of the Ieee 1998;86(11):2278–324. doi: 10.1109/5.726791

18. Farabet C, Couprie C, Najman L, et al. Learning Hierarchical Features for Scene Labeling. Ieee Transactions on Pattern Analysis and Machine Intelligence 2013;35(8):1915–29. doi: 10.1109/tpami.2012.231

19. Krizhevsky A, Sutskever I, Hinton GE. ImageNet Classification with Deep Convolutional Neural Networks. Communications of the Acm 2017;60(6):84–90. doi: 10.1145/3065386

20. Abramoff MD, Folk JC, Han DP, et al. Automated Analysis of Retinal Images for Detection of Referable Diabetic Retinopathy. Jama Ophthalmology 2013;131(3):351–57. doi: 10.1001/jamaophthalmol.2013.1743

21. Deep feature learning for knee cartilage segmentation using a triplanar convolutional neural network. International conference on medical image computing and computer-assisted intervention; 2013. Springer.

22. U-net: Convolutional networks for biomedical image segmentation. International Conference on Medical image computing and computer-assisted intervention; 2015. Springer.

23. LeCun Y, Bengio Y, Hinton G. Deep learning. nature 2015;521(7553):436–44.

24. Deep residual learning for image recognition. Proceedings of the IEEE conference on computer vision and pattern recognition; 2016.

25. Setio AAA, Ciompi F, Litjens G, et al. Pulmonary Nodule Detection in CT Images: False Positive Reduction Using Multi-View Convolutional Networks. Ieee Transactions on Medical Imaging 2016;35(5):1160–69. doi: 10.1109/tmi.2016.2536809

26. Abramoff MD, Lou YY, Erginay A, et al. Improved Automated Detection of Diabetic Retinopathy on a Publicly Available Dataset Through Integration of Deep Learning. Investigative Ophthalmology & Visual Science 2016;57(13):5200–06. doi: 10.1167/iovs.16-19964

27. Esteva A, Kuprel B, Novoa RA, et al. Dermatologist-level classification of skin cancer with deep neural networks. Nature 2017;542(7639):115-+. doi: 10.1038/nature21056

28. Litjens G, Kooi T, Bejnordi BE, et al. A survey on deep learning in medical image analysis. Medical Image Analysis 2017;42:60–88. doi: 10.1016/j.media.2017.07.005

29. van der Heijden AA, Abramoff MD, Verbraak F, et al. Validation of automated screening for referable diabetic retinopathy with the IDx-DR device in the Hoorn Diabetes Care System. Acta Ophthalmologica 2018;96(1):63–68. doi: 10.1111/aos.13613

30. Blondel VD, Guillaume JL, Lambiotte R, et al. Fast unfolding of communities in large networks. Journal of Statistical Mechanics-Theory and Experiment 2008 doi: 10.1088/1742-5468/2008/10/p10008

31. Adai AT, Date SV, Wieland S, et al. LGL: creating a map of protein function with an algorithm for visualizing very large biological networks. Journal of molecular biology 2004;340(1):179–90.

32. Sasaki H, Zhidong L, Sakata I. Academic landscape of hydropower: citation-analysis-based method and its application. International Journal of Energy Technology and Policy 2016;12(1):84–102.

33. Gulshan V, Peng L, Coram M, et al. Development and Validation of a Deep Learning Algorithm for Detection of Diabetic Retinopathy in Retinal Fundus Photographs. Jama-Journal of the American Medical Association 2016;316(22):2402–10. doi: 10.1001/jama.2016.17216

34. Kamnitsas K, Ledig C, Newcombe VFJ, et al. Efficient multi-scale 3D CNN with fully connected CRF for accurate brain lesion segmentation. Medical Image Analysis 2017;36:61–78. doi: 10.1016/j.media.2016.10.004

35. Bejnordi BE, Veta M, van Diest PJ, et al. Diagnostic Assessment of Deep Learning Algorithms for Detection of Lymph Node Metastases in Women With Breast Cancer. Jama-Journal of the American Medical Association 2017;318(22):2199–210. doi: 10.1001/jama.2017.14585

36. Lakhani P, Sundaram B. Deep Learning at Chest Radiography: Automated Classification of Pulmonary Tuberculosis by Using Convolutional Neural Networks. Radiology 2017;284(2):574–82. doi: 10.1148/radiol.2017162326

37. Parmar C, Grossmann P, Bussink J, et al. Machine Learning methods for Quantitative Radiomic Biomarkers. Scientific Reports 2015;5 doi: 10.1038/srep13087

38. Kiranyaz S, Ince T, Gabbouj M. Real-Time Patient-Specific ECG Classification by 1-D Convolutional Neural Networks. Ieee Transactions on Biomedical Engineering 2016;63(3):664–75. doi: 10.1109/tbme.2015.2468589

39. Tabar YR, Halici U. A novel deep learning approach for classification of EEG motor imagery signals. Journal of neural engineering 2016;14(1):016003.

40. Acharya UR, Oh SL, Hagiwara Y, et al. Deep convolutional neural network for the automated detection and diagnosis of seizure using EEG signals. Computers in Biology and Medicine 2018;100:270–78. doi: 10.1016/j.compbiomed.2017.09.017

41. Lin YP, Wang CH, Jung TP, et al. EEG-Based Emotion Recognition in Music Listening. Ieee Transactions on Biomedical Engineering 2010;57(7):1798–806. doi: 10.1109/tbme.2010.2048568

42. Atzori M, Gijsberts A, Castellini C, et al. Electromyography data for non-invasive naturally-controlled robotic hand prostheses. Scientific Data 2014;1 doi: 10.1038/sdata.2014.53

43. Ji SW, Xu W, Yang M, et al. 3D Convolutional Neural Networks for Human Action Recognition. Ieee Transactions on Pattern Analysis and Machine Intelligence 2013;35(1):221–31. doi: 10.1109/tpami.2012.59

44. Ordonez FJ, Roggen D. Deep Convolutional and LSTM Recurrent Neural Networks for Multimodal Wearable Activity Recognition. Sensors 2016;16(1) doi: 10.3390/s16010115

45. Shan CF, Gong SG, McOwan PW. Facial expression recognition based on Local Binary Patterns: A comprehensive study. Image and Vision Computing 2009;27(6):803–16. doi: 10.1016/j.imavis.2008.08.005

46. Begg R, Kamruzzaman J. A machine learning approach for automated recognition of movement patterns using basic, kinetic and kinematic gait data. Journal of Biomechanics 2005;38(3):401–08. doi: 10.1016/j.jbiomech.2004.05.002

47. Tompson J, Stein M, Lecun Y, et al. Real-Time Continuous Pose Recovery of Human Hands Using Convolutional Networks. Acm Transactions on Graphics 2014;33(5) doi: 10.1145/2629500

48. Obermeyer Z, Emanuel EJ. Predicting the Future - Big Data, Machine Learning, and Clinical Medicine. New England Journal of Medicine 2016;375(13):1216–19. doi: 10.1056/NEJMp1606181

49. Uzuner O, South BR, Shen SY, et al. 2010 i2b2/VA challenge on concepts, assertions, and relations in clinical text. Journal of the American Medical Informatics Association 2011;18(5):552–56. doi: 10.1136/amiajnl-2011-000203

50. Mullainathan S, Spiess J. Machine Learning: An Applied Econometric Approach. Journal of Economic Perspectives 2017;31(2):87–106. doi: 10.1257/jep.31.2.87

51. Rajkomar A, Oren E, Chen K, et al. Scalable and accurate deep learning with electronic health records. Npj Digital Medicine 2018;1 doi: 10.1038/s41746-018-0029-1

52. Nemati S, Holder A, Razmi F, et al. An Interpretable Machine Learning Model for Accurate Prediction of Sepsis in the ICU. Critical Care Medicine 2018;46(4):547–53. doi: 10.1097/ccm.0000000000002936

53. Avendi M, Kheradvar A, Jafarkhani H. A combined deep-learning and deformable-model approach to fully automatic segmentation of the left ventricle in cardiac MRI. Medical image analysis 2016;30:108–19.

54. Arbabshirani MR, Plis S, Sui J, et al. Single subject prediction of brain disorders in neuroimaging: Promises and pitfalls. Neuroimage 2017;145:137–65. doi: 10.1016/j.neuroimage.2016.02.079

55. Orru G, Pettersson-Yeo W, Marquand AF, et al. Using Support Vector Machine to identify imaging biomarkers of neurological and psychiatric disease: A critical review. Neuroscience and Biobehavioral Reviews 2012;36(4):1140–52. doi: 10.1016/j.neubiorev.2012.01.004

56. Suk HI, Lee SW, Shen DG, et al. Hierarchical feature representation and multimodal fusion with deep learning for AD/MCI diagnosis. Neuroimage 2014;101:569–82. doi: 10.1016/j.neuroimage.2014.06.077

57. Rathore S, Habes M, Iftikhar MA, et al. A review on neuroimaging-based classification studies and associated feature extraction methods for Alzheimer’s disease and its prodromal stages. Neuroimage 2017;155:530–48. doi: 10.1016/j.neuroimage.2017.03.057

58. Walsh CG, Ribeiro JD, Franklin JC. Predicting Risk of Suicide Attempts Over Time Through Machine Learning. Clinical Psychological Science 2017;5(3):457–69. doi: 10.1177/2167702617691560

59. Heinsfeld AS, Franco AR, Craddock RC, et al. Identification of autism spectrum disorder using deep learning and the ABIDE dataset. Neuroimage-Clinical 2018;17:16–23. doi: 10.1016/j.nicl.2017.08.017

60. Breiman L. Random forests. Machine Learning 2001;45(1):5–32. doi: 10.1023/a:1010933404324

61. Ma JS, Sheridan RP, Liaw A, et al. Deep Neural Nets as a Method for Quantitative Structure-Activity Relationships. Journal of Chemical Information and Modeling 2015;55(2):263–74. doi: 10.1021/ci500747n

62. Alipanahi B, Delong A, Weirauch MT, et al. Predicting the sequence specificities of DNA- and RNA-binding proteins by deep learning. Nature Biotechnology 2015;33(8):831-+. doi: 10.1038/nbt.3300

63. Pedregosa F, Varoquaux G, Gramfort A, et al. Scikit-learn: Machine Learning in Python. Journal of Machine Learning Research 2011;12:2825–30.

64. Zhu XJ, Feng CQ, Lai HY, et al. Predicting protein structural classes for low-similarity sequences by evaluating different features. Knowledge-Based Systems 2019;163:787–93. doi: 10.1016/j.knosys.2018.10.007

65. Cecotti H, Graser A. Convolutional neural networks for P300 detection with application to brain-computer interfaces. IEEE transactions on pattern analysis and machine intelligence 2010;33(3):433–45.

66. Beretta R. A critical review of the Delphi technique. Nurse researcher 1996;3(4):79–89.

67. Ratner M. FDA backs clinician-free AI imaging diagnostic tools. Nature Biotechnology 2018;36(8):673–74. doi: 10.1038/nbt0818-673a

68. Werbos PJ. Beyond Regression: New Tools for Prediction and Analysis in the Behavioral Sciences, 1975.

69. Administration FaD. Implanted brain-computer interface (BCI) devices for patients with paralysis or amputation---Non-clinical testing and clinical considerations. US Food and Drug Administration 2019

70. Takano Y, Kajikawa Y, Ando M. Trends and typology of emerging antenna propagation technologies: Citation network analysis. International Journal of Innovation and Technology Management 2017;14(01):1740005.

